# The Sensation and Pain Rating Scale: easy to use, clear to interpret, and responsive to clinical change

**DOI:** 10.1101/2023.09.08.23295128

**Authors:** Victoria J Madden, Peter Kamerman, Hayley B. Leake, Mark J. Catley, Lauren C. Heathcote, G Lorimer Moseley

## Abstract

**Background:** The Sensation and Pain Rating Scale (SPARS) allows rating of non-painful as well as painful percepts. While it performs well in the experimental context, its clinical utility is untested. This prospective, repeated-measures study mixed qualitative and quantitative methods to examine the utility and performance of the SPARS in a clinical context, and to compare it with the widely used 11-point NRS for pain.

**Methods:** People presenting for outpatient physiotherapy (n = 121) provided ratings on the SPARS and NRS at first consultation, before and after sham and active clinical interventions, and at follow-up consultation. Clinicians (n = 9) reported each scale’s usability and interpretability using Likert-type scales and free text, and answered additional questions with free text. Each data type was initially analysed separately: quantitative data were visualised and the ES II metric was used to estimate SPARS internal responsiveness; qualitative data were analysed with a reflexive inductive thematic approach. Data types were then integrated for triangulation and complementarity.

**Results:** The SPARS was well received and considered easy to use, after initial familiarisation. Clinicians favoured the SPARS over the NRS for clarity of interpretation and inter-rater reliability. SPARS sensitivity to change was good (ESII=0.9; 95%CI: 0.75-1.10). The greater perceptual range of the SPARS was deemed especially relevant in the later phases of recovery, when pain may recede into discomfort that still warrants clinical attention.

**Conclusion:** The SPARS is a promising tool for assessing patient percept, with strong endorsement from clinicians for its clarity and superior perceptual scope.

**Significance:** Clinicians in this mixed-methods study favoured the Sensation and Pain Rating Scale (SPARS) over a 0-10 Numerical Rating Scale for pain because the SPARS provides a clearly labelled range for rating non-painful events, which supports inter-rater reliability and clear interpretation. Clinicians reported rapid adjustment to the SPARS structure. The SPARS had good internal responsiveness to change. The SPARS may be particularly useful as a person recovers from a painful episode, when residual discomfort still requires clinical attention.

## Introduction

In experimental settings, participants are frequently asked to rate the perceived intensity of stimuli across a range of stimulus strengths. Although these stimuli may elicit a range of non-painful percepts, conventional rating scales limit the reporting of non-painful percepts to a single extreme anchor of ‘no pain’. Some investigators have adapted conventional scales using descriptive anchors for certain numbers or ranges for heat stimuli (e.g. 1=non-painful warmth; 2=low pain (Atlas, Bolger, Lindquist, & Wager, 2010), or 1-3=‘heat sensation, not painful’ for heat stimuli (Leandri et al., 2006)), or a ‘pain threshold’ anchor somewhere in the scale’s range (Kunz, Chatelle, Lautenbacher, & Rainville, 2008; Meulders, Vansteenwegen, & Vlaeyen, 2011) for heat or electrical stimuli. Others have used a scale that diverges symmetrically from no sensation to two alternative ‘extreme pain’ anchors for hot or cold thermal stimuli (Morin & Bushnell, 1998). However, many of these scales are stimulus-specific and have undergone limited psychometric testing.

There is a need for a stimulus-agnostic scale that captures a range of percepts from no sensation through to the worst pain imaginable. The Sensation and Pain Rating Scale (SPARS) is an analogue scale with left-most anchor of −50 (‘no sensation’), central anchor of 0 (‘the exact point at which what you feel transitions to pain’), and right-most anchor of +50 (‘the worst pain you can imagine’). Thus, the range from −50 to 0 reflects non-painful percepts; 0 to +50 reflects painful percepts. Experimental testing of the SPARS showed good psychometric properties and, usefully, a stable curvilinear stimulus-response relationship supporting parametric statistics and calculations of percentage changes in pain (in contrast to the conventional NRS or VAS) (Madden et al., 2019; Price, Bush, Long, & Harkins, 1994; Price, McGrath, Rafii, & Buckingham, 1983). Several studies have used (Bedwell et al., 2022; Chaves et al., 2021; Madden et al., 2016; Traxler, Madden, Moseley, & Vlaeyen, 2019) or adapted (Ho et al., 2022) it as an outcome measure.

Although the SPARS was developed for experiments, its broad perceptual range may be useful in the clinical setting. As a person recovers from a painful episode, pain evoked by certain tasks or movements may recede into non-painful percepts that still reflect deviation from a state of ‘fully recovered’. Clinically, these percepts are important in guiding ongoing rehabilitation. For example, reports of stiffness, pulling, non-painful sensitivity to movement, and not feeling ‘quite right’ are all potentially important in pacing return to full loading, or assessing non-tissue-related contributions to pain and disability. Although specific scales exist for certain non-painful percepts (e.g. Westhoff, Buttgereit, Gromnica-Ihle, & Zink, 2008), using a single scale to quantify the overall ‘extent to which the percept deviates from asymptomatic’ may improve clinical reasoning, reporting, and planning. Conventional numerical rating (NRS) and visual analogue scales for pain do not provide this utility. When using a 0-100 scale anchored with at 0 ‘no pain’, a quarter of patients used numbers greater than 0 to rate clinical sensations that they also classified as non-painful (Littman, Walker, & Schneider, 1985). Anecdotally, clinicians report variable NRS thresholds for the transition to pain. This improvised, unstandardised, and idiosyncratic scale use has the potential to obscure clear communication between patient and clinician, between clinicians, and between clinicians and external stakeholders, such as compensation providers.

This mixed-methods study aimed to examine the clinical utility and performance of the SPARS, and compare it to the widely used 11-point (0-10) NRS.

## Methods

### Study overview

This prospective, repeated-measures study was conducted in multiple physiotherapy practices in Australia, with approval from the University of South Australia’s Human Research Ethics Committee (ID: 202696). Data were collected from clinicians and their eligible patients. Figure 1 shows the structure of the study. Data collection comprised two phases that differed only in their scale usage: Phase 1 used both the SPARS and a 0-10 NRS; Phase 2 used only the SPARS. The clinician administered the scale(s) to each participating patient, at six time points: on first presentation of the patient; immediately before and after a therapeutic intervention expected to produce pain relief – an ‘active’ intervention); immediately before and after an intervention’ expected to *not* produce pain relief – a ‘sham’ intervention); and at a follow-up appointment anticipated to be the last consultation (ranged from 5-62 days after first presentation). After collecting and submitting data for both study phases, the clinician completed an online form to provide their perspectives on the SPARS and NRS.

**Figure 1:**
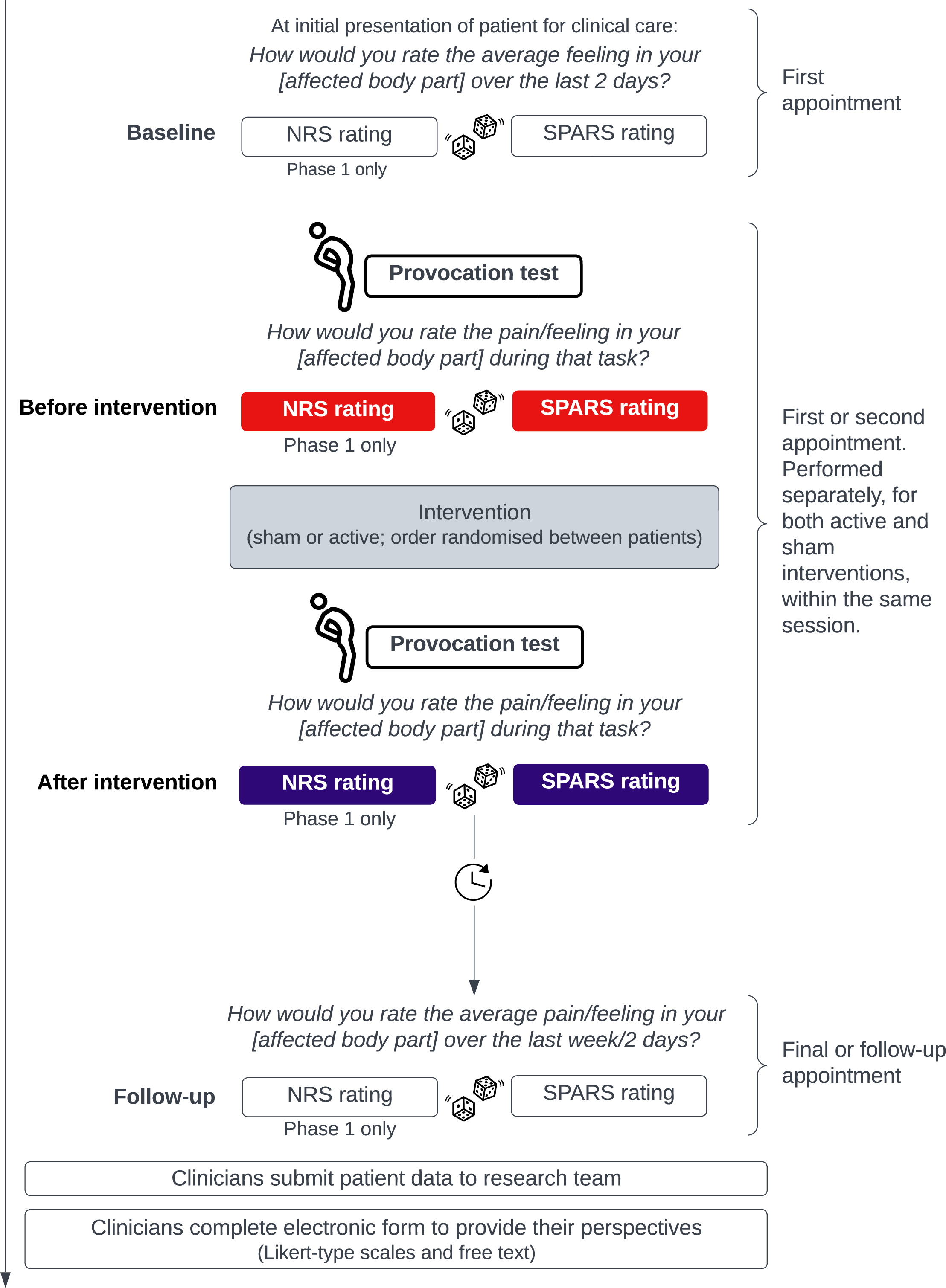
Study structure. NRS ratings were requested in Phase 1only; SPARS ratings were requested in Phases 1 and 2. Dice indicate that the order of assessment was randomised between participants but held constant within each participant. Randomisation was performed by the study team, with MS Excel. Provocation tests and interventions were selected by clinicians, as appropriate to each patient’s presentation. NRS: 0-10 Numerical Rating Scale for pain. SPARS: Sensation and Pain Rating Scale. At the follow-up time point, the phrasing of ‘2days’ was used only if less than one week had passed since the preceding study time point. Icons from the Noun Project: ‘Twist’ by James Keuning; ‘Dice’ by kareemov; ‘Exercise’ by Jeevan Kumar (CC BY 3.0).

We drew on patient ratings to determine SPARS sensitivity to clinical change, to compare the SPARS and NRS for relative coverage of perceptual range and agreement within comparable ranges, and (in light of the historical data referenced above (Littman et al., 1985)) to examine NRS ratings of events also rated as non-painful on the SPARS. We drew on clinicians’ experiences to compare the usability and interpretability of the SPARS and NRS (Likert ratings and free text), to obtain suggestions for improving the utility of the SPARS (free text), and to understand the potential for the SPARS to be used in future clinical practice (free text). We anticipated that, for events rated as painful on the SPARS, agreement between ratings on the SPARS and ratings on the NRS would be good. We anticipated that clinicians would report some difficulty orientating to the SPARS anchors, but that they would favour the non-painful range of the SPARS for its greater coverage of clinically relevant percepts, and that this preference for the SPARS would be more apparent as patients recovered than when patients presented with recent-onset pain.

The study protocol was locked on Open Science Framework (Madden, Kamerman, & Moseley, 2023) (at: https://tinyurl.com/spars-clinical1), after data collection and before data inspection. The protocol specified the intended quantitative analysis but neither the qualitative analysis nor data integration. Supplementary Table S1 fully details the research questions, planned quantitative analysis, notes on any deviations, and the availability of relevant qualitative data. The most important deviations were for blinding of the data analyst (see Blinding section below) and the analytical approach to estimating sensitivity to change.

### Participants

Clinicians were recruited via a clinical advisory network established through the Pain Adelaide Stakeholders’ Consortium, at convenience. Each clinician informed consecutive patients about the study, if they were eligible: aged over 18 years, presenting for treatment from a participating clinician, with English and cognitive proficiency to use each scale (clinician judgement). The target sample sizes (calculations explained in supplementary file) were 96 and 25 patients for Phases 1 and 2, respectively.

### Outcomes

#### Primary outcome – the SPARS

The SPARS was used as a numerical scale. At each assessment, the patient was asked: ‘On a scale from minus 50 to 50, where minus 50 means you feel nothing, where plus 50 is the worst pain possible, and where zero is the exact point at which you feel pain…[phrasing as shown in Figure 1]’.

#### Secondary outcome – 11-point conventional NRS for pain

Phase 1 also used a conventional 11-point NRS for pain, in each of the six conditions described above. The wording for the NRS was: ‘On a scale from 0 to 10, where zero is no pain and 10 is worst possible pain…[phrasing as shown in Figure 1]’.

Descriptive data included patient age, gender (from options: male; female; other), and diagnosis (from options: spinal; limb; pelvic; complex regional pain syndrome; widespread or multiple sites; not listed); order of scale administration (NRS or SPARS first) and interventions (active or sham first), pain provocation test (options: functional; pressure; manual therapy; movement), nature of ‘active’ and ‘sham’ interventions (options: manual therapy; education or advice; an active exercise; active or passive stretch), time between first appointment and follow-up/final appointment (clinician discretion), and patient perspectives (yes/no) at follow-up/final appointment on (a) need for further intervention and (b) whether they considered themselves to have completely recovered.

Clinician feedback included 6 quantitative, Likert-scale questions (response options: very easy, somewhat easy, neither easy nor difficult, somewhat difficult, and very difficult) and 8 qualitative free text questions (full question list in supplementary file). Together, the questions gathered information on ease or difficulty with explaining, using, and interpreting the SPARS/NRS, contextual relevance of the SPARS/NRS, and ideas on ways to make each scale easier to use. They also solicited information on changes over time in clinician confidence and competence with the SPARS, and anticipated future use of the SPARS.

### Manipulations

The clinician chose three manipulations for each patient, according to clinical presentation: a pain provocation test, an ‘active’ intervention, and a ‘sham’ intervention. The pain provocation test was expected to reproduce the patient’s complaint. It could be functional (e.g. sit to stand), movement (e.g. bending the knee), pressure (e.g. firm palpation), or manual therapy (e.g. accessory joint mobilisation). Both interventions were to be brief and plausibly a part of a normal assessment or treatment. The clinician chose an intervention they expected to decrease the patient’s pain for the ‘active’ intervention, and an intervention they expected to have no effect on pain for the ‘sham’ intervention.

#### Blinding

Clinicians were unblinded. Patients were not told the aim of the study, nor the use of active and sham interventions. Data entry assistants were not told the aim of the study, about the range and nature of the SPARS, the order in which assessments were undertaken, or the order of the active and sham interventions. No assessments were undertaken to identify potential loss of blinding. Data analysts were unblinded (protocol deviation 1 of 2; Table S1).

### Data Collection, storage, and analysis

Each clinician and patient was given a unique study ID. Clinicians collected the descriptive and quantitative patient data onto a hard copy form or onto an electronic device (clinician preference) and submitted them via email (to author GLM). Two data entry assistants independently entered clinician-collected data into electronic spreadsheets and compared their entries for accuracy. Clinicians provided quantitative and qualitative data on their perspectives into a single electronic form hosted by SurveyMonkey®. Until data analysis began, clinicians could be re-identified using a separate, password-protected Excel file. Data were tidied and wrangled using R (version 4.2.1 (R. C. Team, 2017)) via RStudio (version 2022.07.0 (R. Team, 2016)) and the following packages: arsenal v. 3.6.3 (Heinzen, 2021), boot v. 1.3.28 (Canty & Ripley, 2021; Davison, 1997), cowplot v. 1.1.1 (Wilke, 2020), epiR v. 2.0.50 (Stevenson & Sergeant, 2022), flextable v. 0.7.2 (Gohel, 2022), ggdist v. 3.2.0 (Kay, 2022), ggpubr v. 0.4.0 (Kassambara, 2020), Gmisc v. 3.0.0 (Gordon, 2022), gridExtra v. 2.3 (Auguie, 2017), janitor v. 2.1.0 (Firke, 2021), kableExtra v. 1.3.4 (Zhu, 2021), knitr v. 1.39 (Y. Xie, 2014, 2015, 2022), lme4 v. 1.1.29 (Bates, 2015), patchwork v. 1.1.1 (Pedersen, 2020), rmarkdown v. 2.14 (Allaire, 2022; Y. Xie, Christophe Dervieux, and Emily Riederer, 2020; Y. Xie, J. J. Allaire, and Garrett Grolemund, 2018), tidyquant v. 1.0.4 (Dancho, 2022), tidyverse v. 1.3.1 (Wickham et al., 2019), and grateful (Rodríguez-Sánchez, Jackson, & Hutchins, 2023).

### Data analysis

This study used both quantitative and qualitative data with the goal of addressing the research questions in a convergent mixed methods design. We mixed the two methods in design, data collection, in a process of integration after initially separate analysis, and in interpretation. Although most of the research questions (Table S1) were designed for specific data types, we had planned to use any available data, even if it provided information for a question for which it was not originally intended. For example, if the qualitative data unexpectedly provided comments on sensitivity to change, we planned to present those data alongside the quantitative data. The data analysis script output can be found at (Madden et al., 2023) (https://tinyurl.com/spars-clinical1). Here we present data visually and in tables. In the tables, unless otherwise specified, p-values are based on a two-sample t-test (for continuous variables) or a chi-square (for categorical variables), as per the ‘arsenal’ package (Heinzen, 2021) for R.

### Quantitative analyses

Although our protocol had locked in preliminary plans for quantitative analysis, subsequent discussion raised the need to revise the approach for assessing sensitivity to change, given the lack of a suitable external reference for the perceptual range covered by the SPARS. We therefore focused on internal responsiveness (protocol deviation 2 of 2). We visualised the change in SPARS before and after the active vs the sham intervention and calculated the ‘effective size II’ (ES II) metric (also ‘standardised response mean’ or ‘responsiveness-treatment coefficient’ or ‘efficiency index’). ES II estimates the change represented by the measure, standardised to the between-participant variability in the change scores, and is not dependent on sample size (Husted, Cook, Farewell, & Gladman, 2000). An ES II value greater than 0.80 has been suggested to represent large responsiveness, yet confidence intervals may more useful than benchmarks for supporting interpretation, so we also generated bias-corrected and accelerated bootstrap confidence intervals. To compare coverage of perceptual range between the SPARS and NRS, we visually compared the ranges of ratings within each condition, between the two scales.

To assess agreement between SPARS and NRS ratings within comparable ranges, we selected only those events that had been rated between 0 and 50 on the SPARS and then divided SPARS ratings by 5. We visualised the data with Bland-Altman plots and a scatterplot overlain with the best-fit regression line and its 95% confidence interval (dependent variable: SPARS; independent variable: NRS). We quantified agreement using Lin’s concordance correlation coefficient (CCC) (Lawrence, 1989, 2000), which is suited to two methods of measuring the same continuous variable (Watson & Petrie, 2010). Lin’s CCC has a maximum value of 1, which indicates perfect agreement. To understand how patients used the NRS to rate non-painful events, we selected only the events rated below 0 on the SPARS and visualised the SPARS and NRS ratings for those events. As an internal check of the study method, we investigated whether the task of giving rating on an NRS influenced SPARS ratings. To do this, we visually compared the central tendency and distributions of SPARS ratings across the two study phases, given that NRS ratings were requested in Phase 1, but not in Phase 2.

### Qualitative data analysis

Qualitative data were analysed using reflexive thematic analysis (Braun & Clarke, 2021). We used an inductive approach in which all the available data were coded, and the codes and resultant themes were developed based on the raw data without an *a priori* coding system. The first author (VJM) familiarised herself with the data by reading and re-reading, and recursively coding the data. Codes reflected semantic and latent features of the data, to capture both literal meanings and contextual interpretations of what participants said and how they said it. Codes were reviewed and refined via discussions between VJM and HBL, to generate potential and then final themes and to name themes. For context, VJM is a clinical and research physiotherapist with little experience in qualitative data analysis but over 10 years’ experience in clinical practice. VJM led the initial development and first experimental testing of the SPARS. HBL is a clinical and research physiotherapist who has led and published mixed methods studies. Neither VJM nor HBL was involved in data collection or in the clinical consultations. The full qualitative dataset is provided in the analysis script output at (Madden et al., 2023)(https://tinyurl.com/spars-clinical1).

### Integration of data

Integration of data allows for deeper insights and capturing nuances in participants’ responses that might not be revealed by only one data type. We used the qualitative data to corroborate, explain, illustrate, and add nuance to the quantitative results, especially regarding problems and proposed solutions. This broadly aligns with the Bryman (2006) justifications of triangulation (corroboration/confirmation) and complementarity (clarification of results from one method by results from another, or expansion). Data were integrated after initial, separate analysis. Where more than one data type shed light on the same topic, the data have been presented using a joint display to identify points of divergence and convergence between the two data types.

## Results

### Participants

The respondent sample included nine treating clinicians (4M, 5F, average (range) of years practising = 8 (3 – 31)) collectively reporting data from a total of 121 patients (Table 1). There was one SPARS rating of 51 (which falls outside the range of the scale), which we retained for analysis on the assumption that the patient had indeed given that rating. The most common diagnosis reported was spinal pain (n = 50), followed by limb pain (n = 30) (Table 1). Each clinician reported data from between 23 (clinician 1) and three (clinician 9) patients (Table S2). There were 97 patients included in Phase 1, representing 464 rating pairs for painful events (minimum sample size = 87 rating pairs), and 24 patients included in Phase 2 (target sample size = 25).

**Table 1:** Patient information by study phase. CRPS: Complex regional pain syndrome.

|  | phase 1 (N=97) | phase 2 (N=24) | Total (N=121) |
| --- | --- | --- | --- |
| **Gender** |  |  |  |
| Female | 45 (46.4%) | 17 (70.8%) | 62 (51.2%) |
| Male | 48 (49.5%) | 7 (29.2%) | 55 (45.5%) |
| Other | 4 (4.1%) | 0 (0.0%) | 4 (3.3%) |
| **Age (yrs)** |  |  |  |
| Mean (SD) | 41 (10) | 40 (9) | 41 (10) |
| Range | 18 - 66 | 22 - 64 | 18 - 66 |
| **Diagnosis** |  |  |  |
| CRPS | 7 (7.2%) | 2 (8.3%) | 9 (7.4%) |
| Limb | 26 (26.8%) | 4 (16.7%) | 30 (24.8%) |
| Pelvic | 7 (7.2%) | 2 (8.3%) | 9 (7.4%) |
| Spinal | 38 (39.2%) | 12 (50.0%) | 50 (41.3%) |
| Widespread | 19 (19.6%) | 4 (16.7%) | 23 (19.0%) |

The most common provocation tests, ‘active’ interventions, and ‘sham’ interventions chosen by clinicians were functional (chosen 71 times), education/advice (reported 50 times), and manual therapy (reported 26 times), respectively (Table 2).

**Table 2:** Interventions used and need for ongoing treatment.

|  | Overall (N=121) |
| --- | --- |
| **Order in which scales were used** |  |
| NRS then SPARS | 61 (50.4%) |
| SPARS then NRS | 60 (49.6%) |
| **Provocation test** |  |
| functional | 71 (58.7%) |
| manual therapy | 11 (9.1%) |
| movement | 25 (20.7%) |
| pressure | 14 (11.6%) |
| **Active intervention** |  |
| N missing | 17 |
| active exercise | 28 (26.9%) |
| education/advice | 50 (48.1%) |
| manual therapy | 26 (25.0%) |
| **Sham intervention** |  |
| N missing | 25 |
| active exercise | 26 (27.1%) |
| education/advice | 25 (26.0%) |
| manual therapy | 45 (46.9%) |
| **Days from baseline to follow-up time point** |  |
| Mean (SD) | 32 (14) |
| Median | 30 |
| 25th, 75th percentile | 22, 42 |
| Range | 5 - 62 |
| **Do you think you still need treatment?** |  |
| no | 44 (36.4%) |
| yes | 77 (63.6%) |
| **Do you think you have completed your recovery?** |  |
| no | 107 (88.4%) |
| yes | 14 (11.6%) |

The median (interquartile range) period from baseline ratings to follow-up ratings was 30 (22-42) days. At follow-up, 64% of patients (n = 77) said they still needed treatment. Those who reported still needing treatment had a marginally *longer* period to follow-up assessment, higher SPARS and NRS ratings for baseline pain and follow-up pain, and a smaller reduction in SPARS ratings but a counter-intuitively *greater* reduction in NRS from baseline to follow-up, compared to those who said they no longer needed treatment (Table S3). Also at follow-up, 88% (n = 107) reported they had not completely recovered. This group had higher SPARS and NRS ratings for baseline pain and follow-up pain, and a smaller reduction in ratings from baseline to follow-up in both scales, than those who said they had completely recovered (Table S4).

### Quantitative results

Figure 2 shows clinicians’ responses to the six questions asked about the SPARS and NRS. All the clinicians rated explaining, reporting, and interpreting the SPARS as either very easy or easy. All clinicians also rated explaining the NRS as ‘very easy’ or ‘easy’, but one gave an judgement of how easy it was for patients to report an NRS rating, and three clinicians reported that it was difficult to interpret their patients’ NRS ratings. Overall, these data suggest the SPARS may support greater interpretational clarity than the NRS.

**Figure 2:**
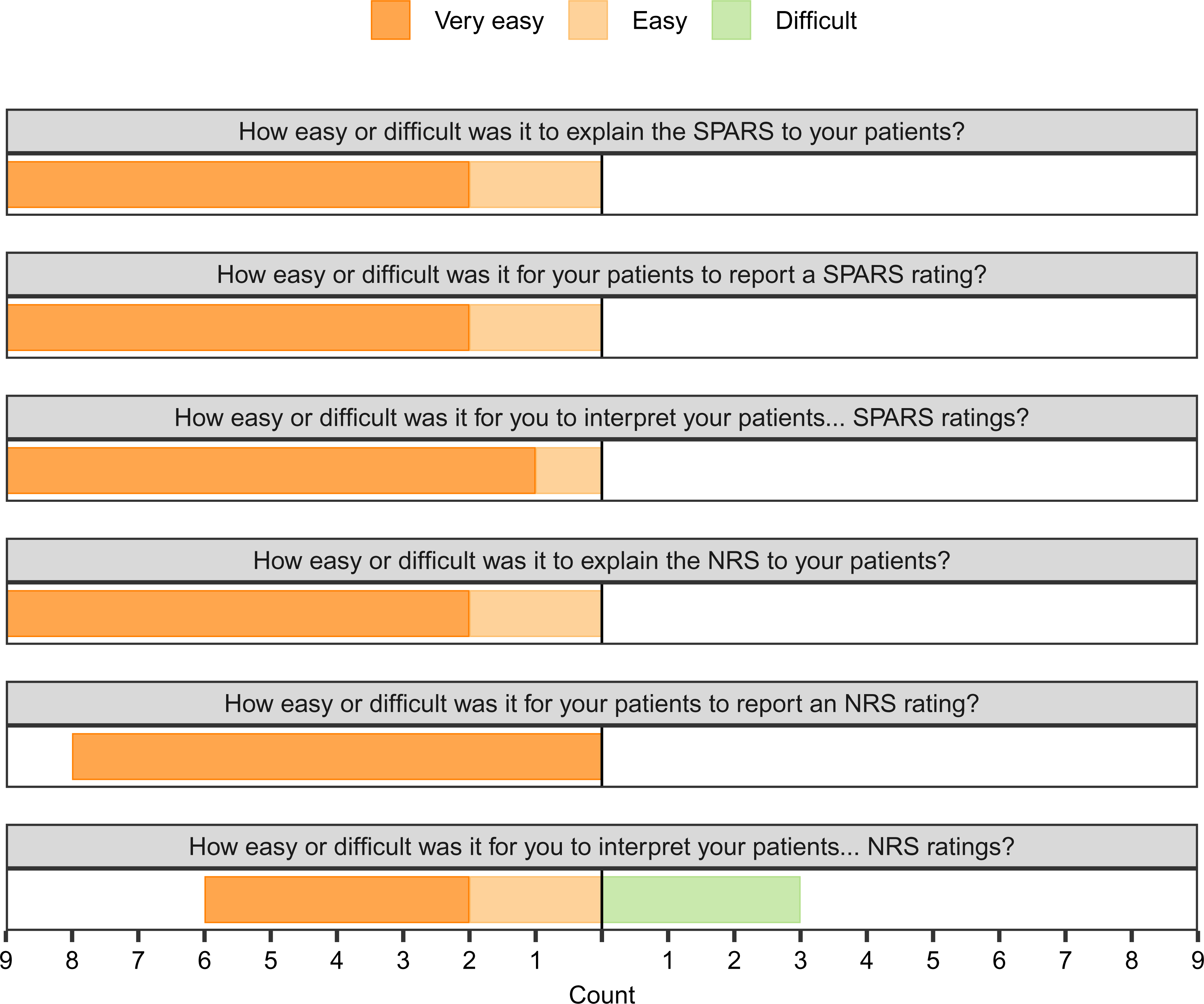
Clinician reports on explaining, using, and interpreting the SPARS and NRS; data coloured by response. Endorsements of ‘Neither easy nor difficult’ are not shown. Response option ‘Very difficult’ was never endorsed.

The responsiveness of the SPARS to clinical change elicited by an active intervention is shown in Figure 3 and represented by the ES II statistic. Figure 3 shows some spread of ratings into the non-painful range of the SPARS after active intervention that is not seen after sham intervention. The ES II estimate was 0.9 (95% CI: 0.75 – 1.10), suggesting excellent internal responsiveness to change.

**Figure 3:**
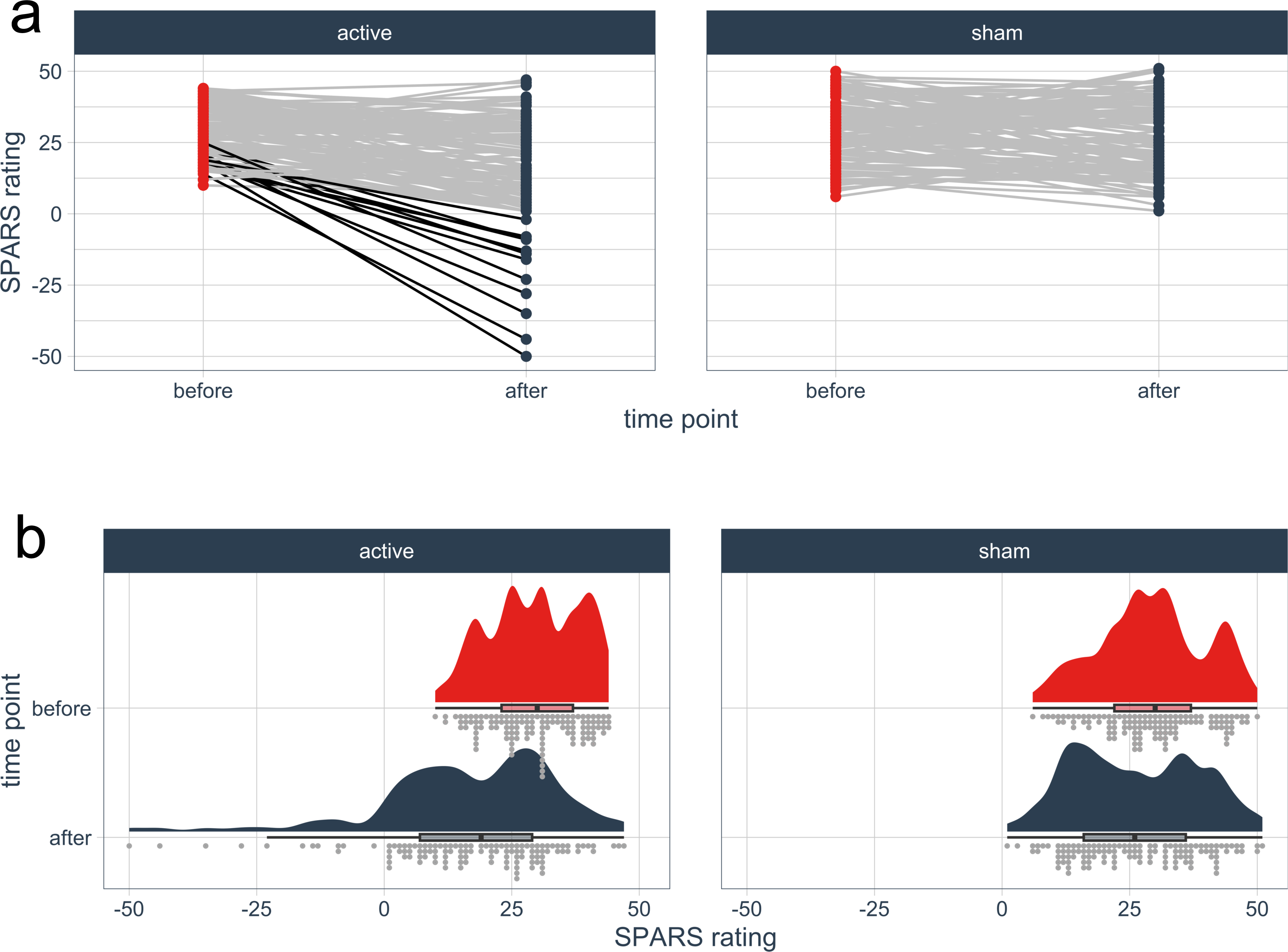
Change in SPARS ratings with intervention. a) Change in ratings by individual patients. Each line represents the change in ratings from one patient; each patient underwent both the active and sham interventions. Lines are darkened to improve visibility of data for patients who reported a SPARS rating < 0. b) Tukey boxplot shows median, two quartiles, and extremes after excluding outliers as per the ggdist package. Each rating is represented by one dot.

Figure 4 shows the range of each scale that was used when the SPARS and NRS were used to rate the same event. Within the working range of the NRS (0 to 10), and the pain range of the SPARS (0 to 50), there was a linear relationship between SPARS and NRS ratings. However, this relationship broke down for the ‘after active intervention’ and at ‘follow-up’ situations, where the NRS reached a minimum value of 0 and provided no further values for assessing non-painful events. The SPARS did not have this lower limit, and continued to allow ratings of sensations below pain threshold (0 to −50).

**Figure 4:**
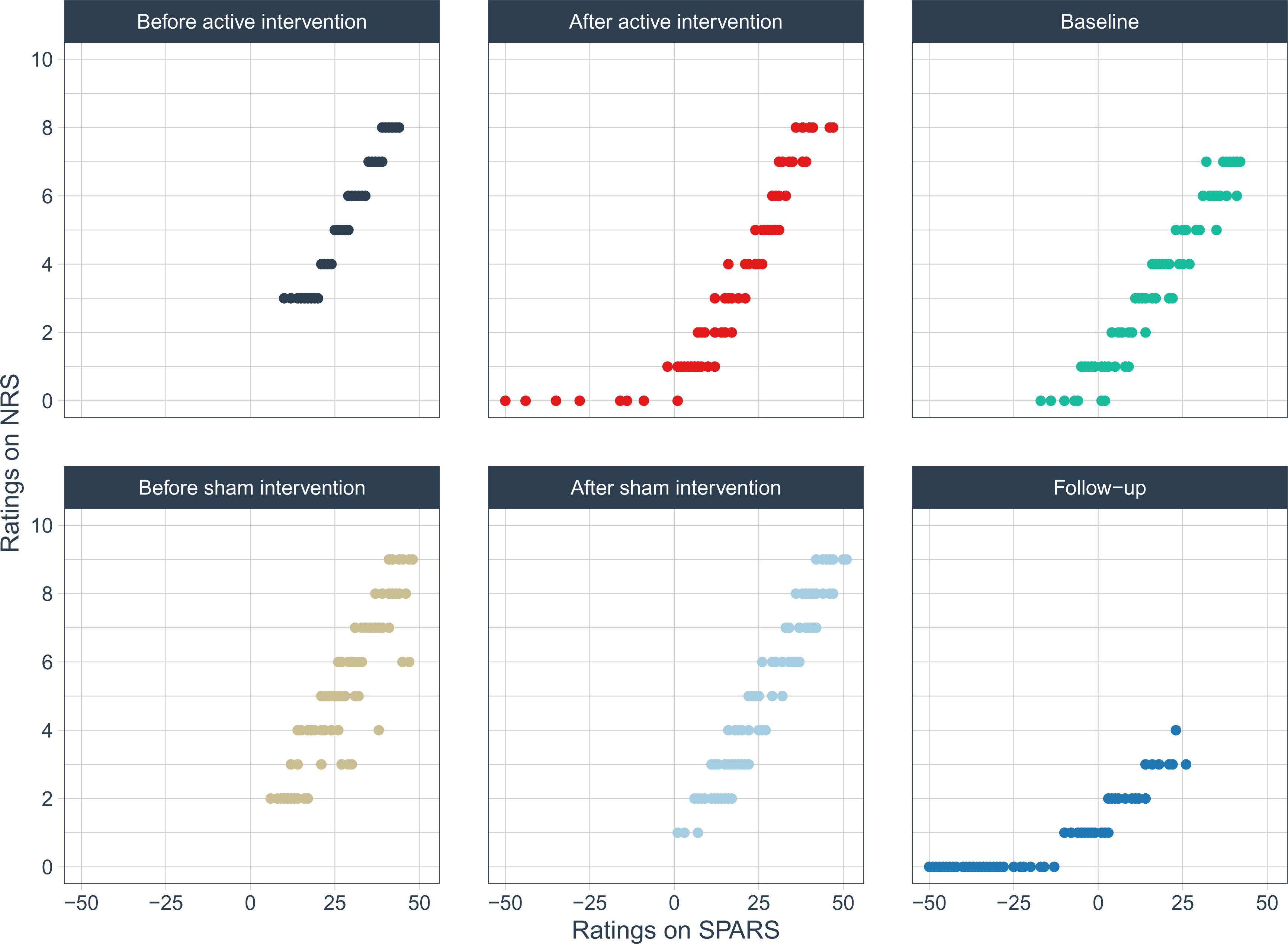
Ratings on SPARS vs ratings on NRS, faceted and coloured by the condition under which ratings were given (582 paired ratings across 6 conditions).

Figure 5 addresses agreement between the two scales, in comparable ranges. The Bland-Altman plots in indicate that the average difference between measurements was less than 5% of the range (which was 0-10) for all conditions. That the 95% CI for the slope of the regression line included 1 in all 5 conditions with sufficient data suggests good agreement. Table 3 shows concordance correlation coefficient values for the same data, by condition. Although the rules of thumb for interpreting CCC are variable and controversial, none of the values found was concerning.

**Figure 5:**
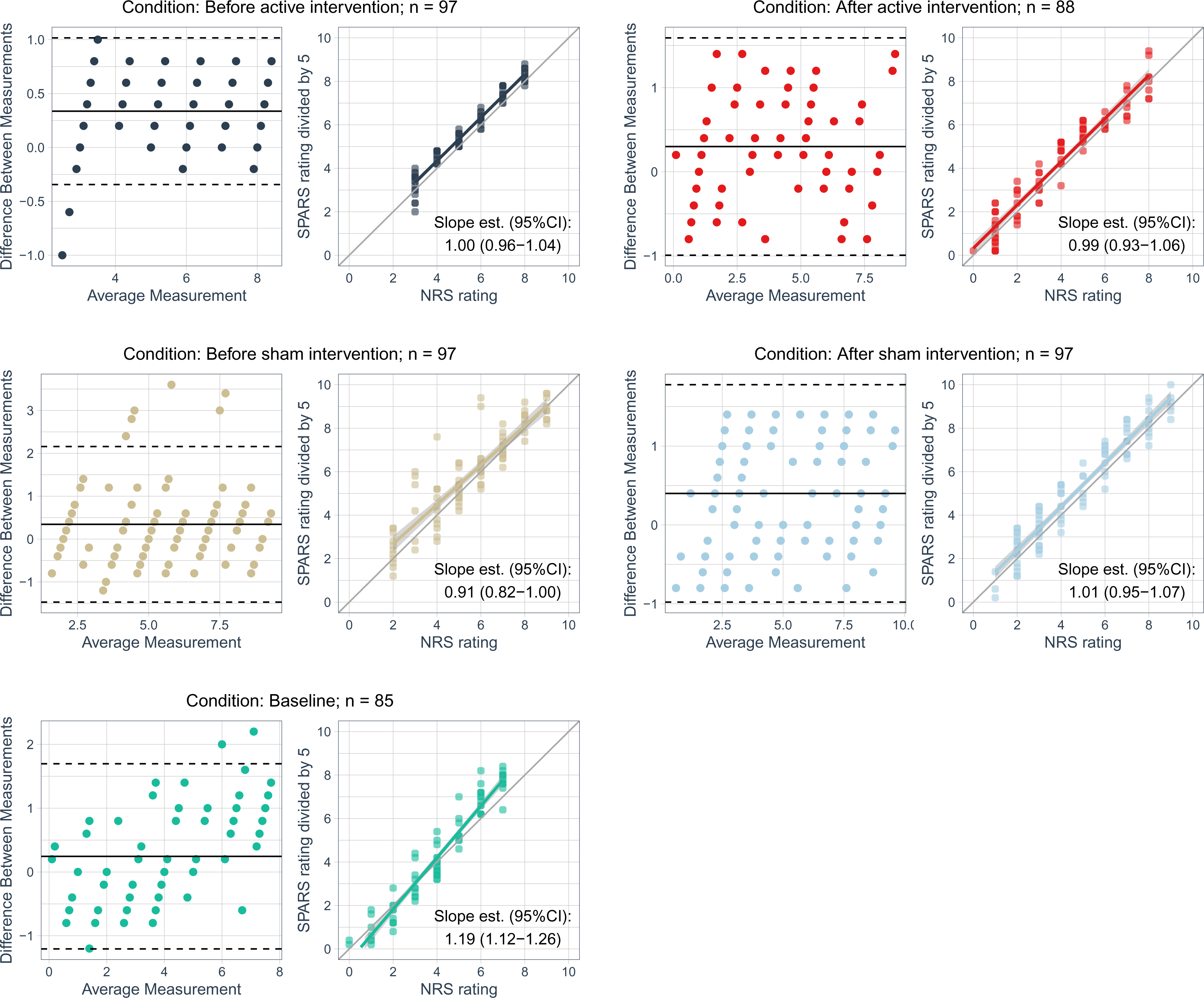
Bland-Altman (left) and regression (right) plots of events rated SPARS ≥ 0, faceted by condition. There were too few data points (n = 25) to provide a useful analysis for the condition of 1-week average at follow-up. Data points are coloured by condition. For the Bland-Altman plots, the black solid (stippled) line: average (95% CI) difference in measurements. For the regression plots: i) the solid coloured line shows the line of best fit, ii) the grey line is the line of identity, and iii) Slope est. (95% CI) is the beta coefficient, with 95% confidence interval, for the regression line of best fit.

**Table 3:** Concordance correlation coefficients (CCC) for each condition when SPARS data were divided by 5.

| Condition | CCC (95% CI) |
| --- | --- |
| Before active intervention | 0.96 (0.95 - 0.97) |
| After active intervention | 0.95 (0.93 - 0.97) |
| Before sham intervention | 0.89 (0.84 - 0.92) |
| After sham intervention | 0.95 (0.92 - 0.96) |
| Baseline | 0.94 (0.92 - 0.96) |

#### Use of NRS for ‘non-painful’ events

Figure 6 shows ratings on the SPARS and NRS for events that were rated in the non-painful range of the SPARS, and that were also rated on the NRS. Only three of the six conditions had events that satisfied these criteria. NRS ratings ranged from 0 to 1; SPARS ratings ranged from −50 to −1.

**Figure 6:**
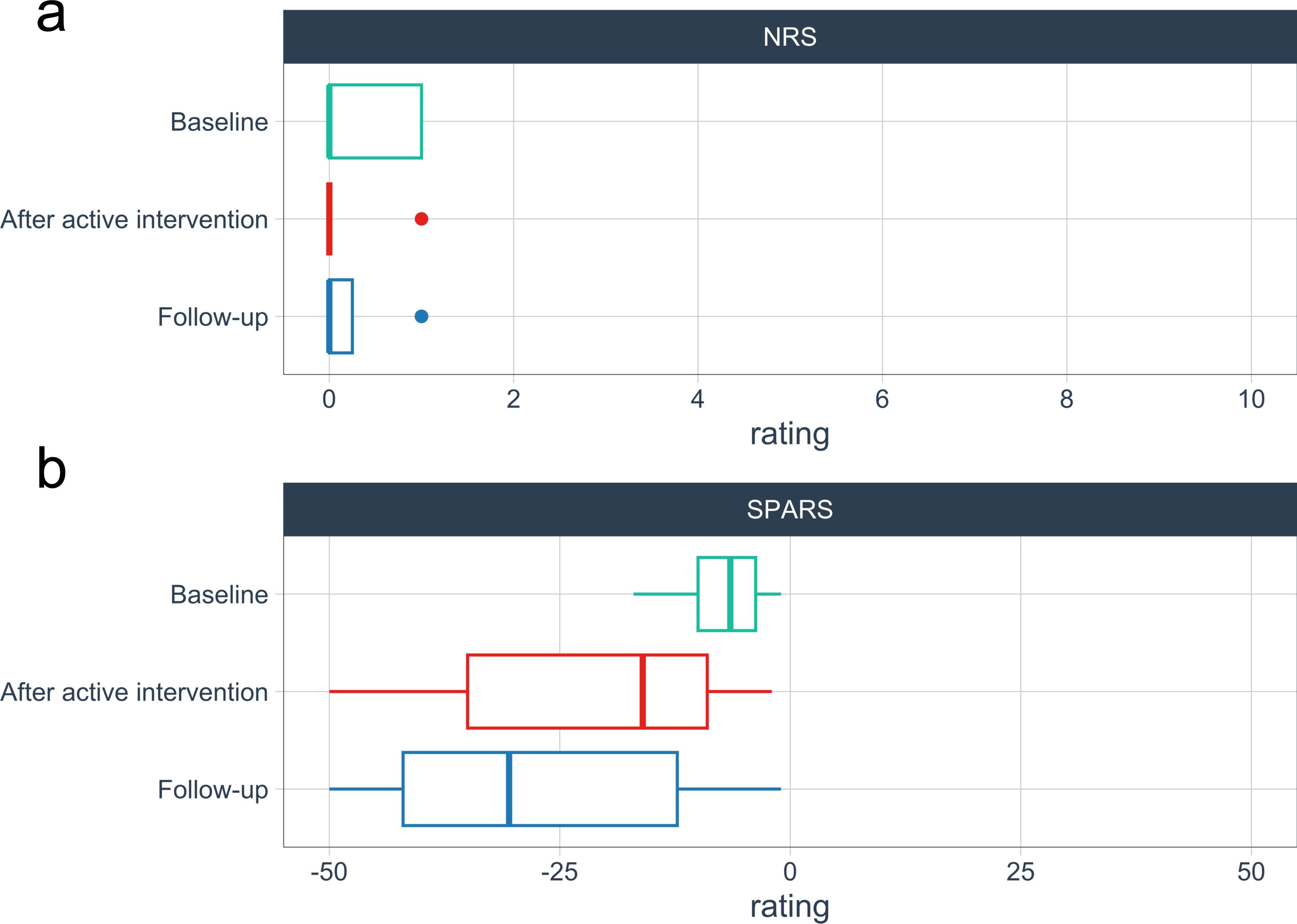
Distribution of ratings of events rated in ‘non-painful’ range of the SPARS (SPARS < 0), faceted by scale. Average over last week, at follow-up: 72 non-painful events of 97 (74% of total events in this condition); Average over 2 days, at baseline: 12 non-painful events of 97 (12%); After active intervention: 9 non-painful events of 97 (9%).

Figures 7 and 8 display the central tendency and distribution of SPARS ratings by study phase, to clarify whether SPARS ratings were affected by collecting NRS ratings. Although visual inspection of Figure 8 suggests greater spread of SPARS ratings after the active intervention in Phase 1 than in Phase 2 (middle right facet), this was not statistically significant.

**Figure 7:**
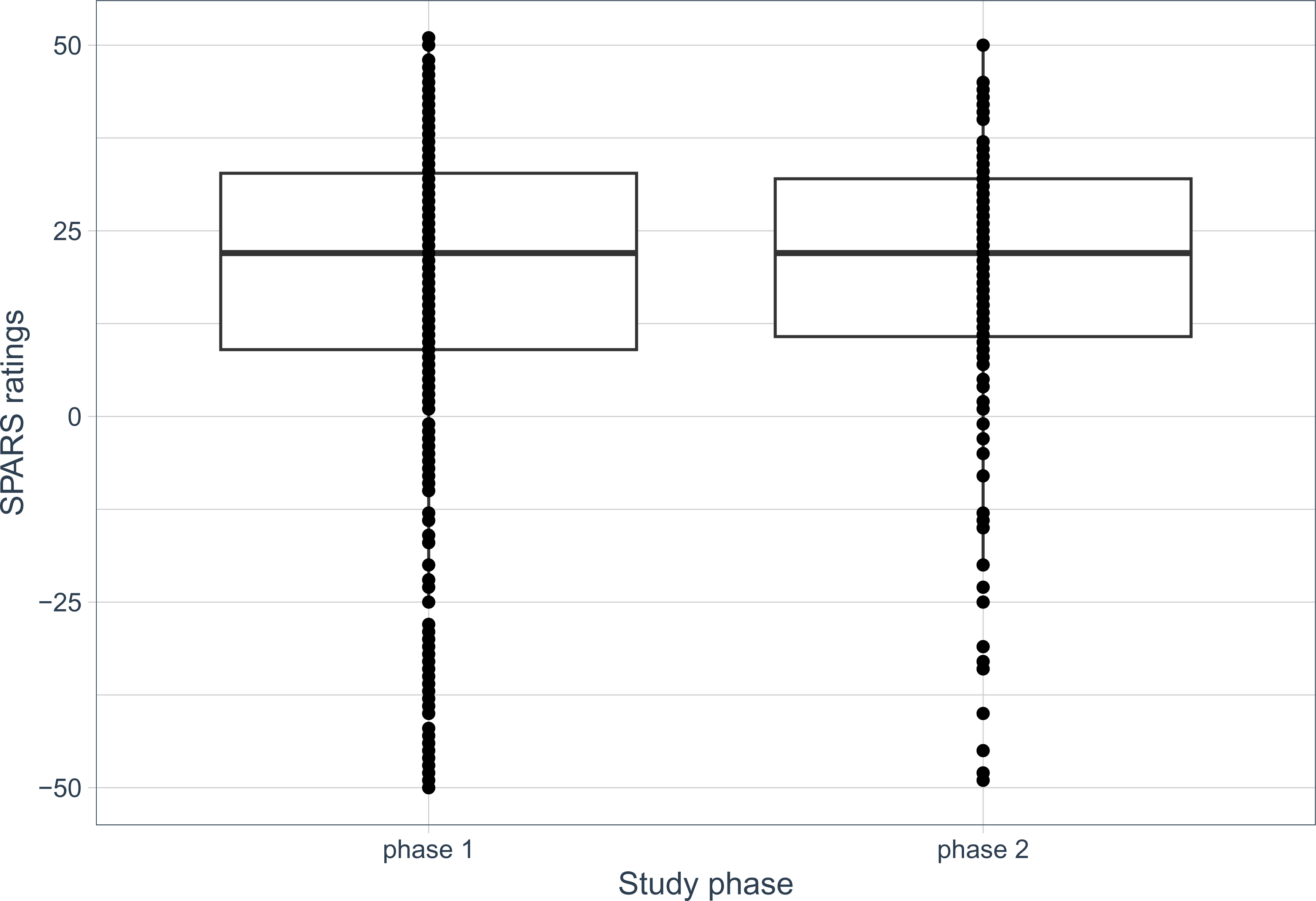
SPARS ratings by study phase. Each dot represents one rating. Boxplot shows median and interquartile range.

**Figure 8:**
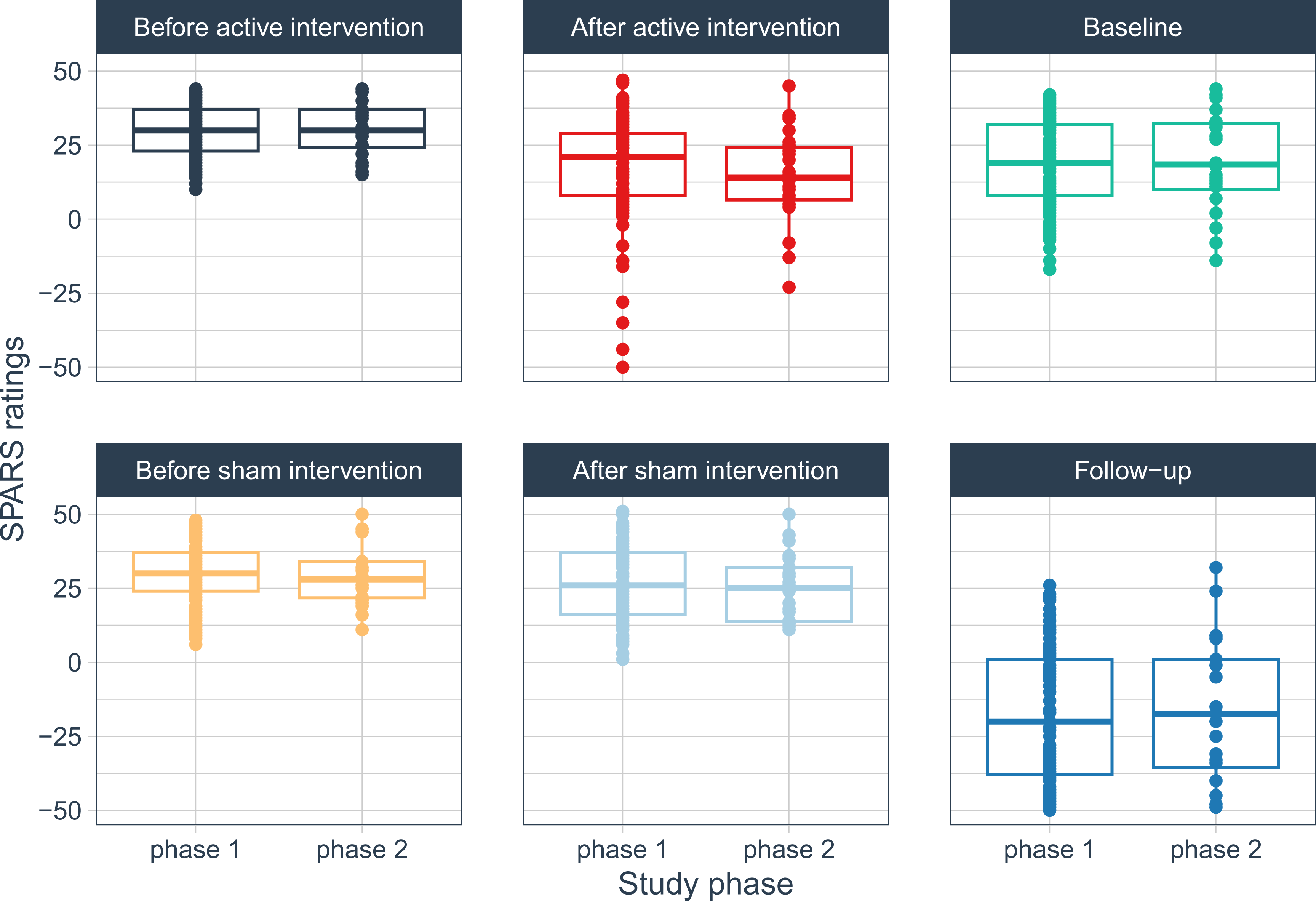
SPARS ratings by study phase, faceted and coloured by condition under which ratings were given. Each dot represents one rating. Boxplot shows median and interquartile range.

### Qualitative results

We generated two overarching themes related to clinicians’ experiences using the SPARS: (1) Easy to use after brief familiarisation, and (2) Avoids several pitfalls of the NRS.

#### Theme 1: The SPARS was easy to use after brief familiarisation

Overall, clinicians reported positive experiences using the SPARS. Although some clinicians reported an adjustment period from conventional scales to the SPARS (“*they take a little while to get the hang of the new scale”, 42)*, the adjustment was quick (“*All my patients got the hang of it quickly”, 32)*. To help patients adjust to the SPARS, clinicians reported that careful explanation of the SPARS structure was necessary (“*The main thing for me was the initial explanation”, 29)*. Most clinicians expressed that, after a practice period, the SPARS structure was easy enough to support rapid adjustment (“*it was very quick - a couple of run throughs and then the wording was easy and I am confident now”, 69)*.

Some clinicians reported challenges adjusting to the SPARS, irrespective of the speed of adjustment (“*I found it difficult at first to adjust to the SPARS”, 42)*. Some of these challenges stemmed from the deeply ingrained familiarity with the previous pain rating scale – the NRS. Other challenges were attributed to the perceived lack of intuitiveness of the SPARS, in comparison to the NRS (“*initially it was not as intuitive as the 0 - 10 scale”, 32)*. The difference between the perception of intuitiveness between the SPARS and NRS appeared to reflect discomfort with the numbering: the two ranges of the SPARS (which are delineated by the SPARS anchors) had to be clarified for patients (*“once patients understand that it is a −50 to 50 scale and that pain starts at 0, then it is fine”, 46)*. Visual and electronic aids were proposed to facilitate understanding and use of the SPARS. Specifically, clinicians suggested creating an educational wall poster (“*a nice-looking infographic “, 42)* or a website to serve as a convenient reference for clinicians when communicating with patients. Others suggested adapting the SPARS as an electronic version, to smooth the familiarisation processes. Regardless of format, clearly written instructions were recommended (*“better instructions above the scale”, 69*).

One clinician requested web-based access to a comprehensive explanation and comparative data on the SPARS, to prevent clinicians from having to make assumptions about how SPARS ratings relate to ratings on other scales (“*I am just presuming that 25 on the SPARS matches 5 on the NRS, but is that how it really works?”, 32)*. The lower anchor of the SPARS was also flagged as needing clarification: *“I had a few occasions when I was unsure how to define the [-50] end of the SPARS because feeling ‘nothing’ wasn’t quite right. I used ‘completely normal’ for one patient but that wouldn’t be very good for wider use because ‘completely normal’ might still be uncomfortable or even painful”, 69).* Clinicians liked that the SPARS allowed patients to describe sensations that were *“not painful but don’t feel right” (29)*. This was considered useful for tracking progress after pain had resolved (*“…it allowed [patients] to communicate the feeling that there was still some improvement to go even though they weren’t really having pain anymore”, 29)*.

On the whole, this theme captured generally positive feelings towards the SPARS, a brief period of adjustment, and some possible tools to support implementation of the SPARS in the clinical setting.

#### Theme 2: The SPARS avoids several pitfalls of the NRS

The second theme captures clinicians’ reports that the SPARS offers a greater utility and interpretational clarity than the conventional NRS. Several clinicians noted concerns about the reliability of the NRS, and said they routinely added *ad hoc* anchors to the NRS to clarify a point of transition to pain. They noted that this added anchor was not necessary with the SPARS. *“[on the NRS] when people rate their pain as 1 or 2 and when I question them about it they actually say it is not so much painful as uncomfortable. I can explain it but then the ratings are dependent on me clearly talking them through 0 to 2 … we… nearly decided that we would say that 2 out of 10 was when you are definitely sure it is painful. I think the SPARS solves this problem and I really like it. We all like it here.“ (69)* This lack of clarity was problematic enough for one clinic to have developed a “policy” to manage it, that even had to be explained to external stakeholders: *“The primary implementation strategy that we use to facilitate interpretation and indeed utility of the NRS is to make 1 equal to the point at which it is definitely painful. We tend to articulate that as the pain threshold. I suspect it would assist us and our colleagues if [using SPARS] was a formal modification. This way we would no longer be compelled to provide an explanation of our policy when dealing with external stakeholders. (21)*

Clinicians reported that using the SPARS had revealed previously unidentified problems with inter-rater reliability of the NRS linked to variable explanations for the lower numbers on the NRS: *“Some [clinicians] were telling patients that 1 was more discomfort than pain and some were saying that 2 was when it was definitely painful… if the person came back to see someone else, they may score the same thing differently on the normal pain scale, but not on the SPARS.” (25)* Clinicians described the SPARS as a superior tool because it resolved problems of inter-rater reliability by offering distinct categories of “no pain” and “no sensation”. *“I think this is where the SPARS has a clear advantage. [With] the VAS [visual analogue scale] for pain… we still get questions such as ‘is this point (pointing to the left-hand end) where I feel absolutely nothing or is it where I feel some discomfort or stiffness?’” (08).* Some participants even suggested a formal modification for the NRS, to improve interpretational clarity, by “*label[ling] 1 as something akin to the zero point on the [SPARS].” (21)*

Clinicians viewed the SPARS as better suited than the NRS to capturing patients’ clinical ratings that transitioned through a range of sensations from no sensation, through a non-painful range, into a painful range. This suggests that the SPARS was thought to have greater face validity than the NRS. *“This is where I think the SPARS is better than our other methods of assessing pain to be honest with you… you don’t have no sensation at all and then suddenly pain and I know some patients who talk about being sore but not painful for example” (46).* This greater face validity translated to a reminder that patients’ treatment goals go beyond just reducing pain*: “it is just a reminder that their treatment goals go beyond just getting pain down; almost aiming higher sort of this?” (32)*

Overall, this second theme captures that clinicians considered the SPARS to address known problems with the NRS, inferring that the SPARS was superior to the NRS. The superior utility and interpretational clarity of the SPARS was attributed to its anchors and its explicit labelling of a pain-free range.

Together, these two themes appeared to culminate in a notable enthusiasm for ongoing use of the SPARS, especially later in recovery: *“It is especially helpful when patients are getting better but still feel like they have a way to go to be ‘just right’" (25)*. Several clinicians reported they now encourage the use of SPARS in their clinic, or have implemented it and even encouraged other clinicians to use it. However, the potential for broad use of the SPARS was limited by the scales required by official reports, and a reluctance to have two pain scales in use: *"we still have to use the NRS too for some reports and I don’t think we will use both.” (69)*.

### Integration of results

There were four topics on which both quantitative and qualitative data were available. Table 4 provides a joint visual display of quantitative and qualitative data by topic.

**Table 4:** Joint visual display of quantitative, qualitative, and mixed-methods meta-inferences by topic.

| Topic | Quantitative result | Qualitative excerpts | Meta-inference about complementarity |
| --- | --- | --- | --- |
| Ease of explanation | Both the SPARS and NRS were rated as easy or very easy to explain | "[The SPARS] is very easy to explain" (46)<br>"I found it difficult at first to adjust to the SPARS because I am so used to using the zero to ten scale" (42)<br>"I think they are equally easy to use" (08)<br>"Really [the SPARS] is no harder to use than the usual NRS, just less familiar." (29) | *Expansion*<br>Clinicians described an initial period of familiarisation before use of the SPARS was easy; this was not identified in the Likert scale ratings. |
| Ease of reporting | All clinicians but one reported that it was very easy or easy for patients to report on the SPARS or the NRS; one clinician provided an ambiguous response for the NRS. | [On the SPARS:] "I think the explanation is easy and patients get it" (46)<br>"All my patients got the hang of [the SPARS] quickly but initially it was not as intuitive as the 0 - 10 scale (32)<br>"Once they understand that it is only positive numbers that are about pain then [the SPARS] is easy as pie" (29) | *Expansion*<br>Patients needed familiarisation before reporting on the SPARS was easy; this was not identified in the Likert scale ratings. |
| Ease of interpreting | The SPARS was easy or very easy to interpret. In contrast, the NRS was noted as difficult to interpret by three of the nine clinicians (25, 42, 69), whereas the remaining clinicians rated it as easy or very easy to interpret. | "[The SPARS] is way more informative to us" (42)<br>"[Interpretation] is where I think the SPARS is better than our other methods of assessing pain" (46)<br>"The SPARS is much easier... because it makes very clear where the pain threshold is" (42)<br>"[On the SPARS] zero really was zero and it was particularly interesting when patients were about zero and were really evaluating things" (29) | *Confirmation*<br>Interpreting the SPARS was viewed as easier than interpreting the NRS and other conventional assessments of pain. |
| Does the SPARS provide quantification over a greater perceptual range than a 0–10 Pain NRS? | NRS scores transformed to a 100-point scale had greater spread than SPARS scores in 4 of the 6 conditions. However, the largest difference in score range was for average pain rating over the week preceding the follow-up consultation, where SPARS ratings covered more than twice the range of transformed NRS ratings. | Allows patients to express sensation that is 'not quite right', although not painful:<br>*"This is where I think the SPARS is better than our other methods of assessing pain to be honest with you... you don't have no sensation at all and then suddenly pain, and I know some patients who talk about being sore but not painful, for example."* (46)<br>*"...[the SPARS] allowed them to communicate the feeling that there was still some improvement to go even though they weren't really having pain anymore."* (29)<br>*"I think [the SPARS] is more useful as people get better"* (29) | *Expansion*<br>The non-painful range of the SPARS may be most valuable during the later stages of recovery from a painful episode, when its ability to capture percepts that are non-painful but also not 'normal', may be clinically informative. |

## Discussion

This prospective study mixed quantitative and qualitative data to examine the utility and performance of the SPARS, and to compare the SPARS with the conventional 0-10 NRS for pain, in a clinical context. Overall, participant feedback on the SPARS was positive. Clinicians described the SPARS as “conceptually attractive” and reported that it was easy to use after initial familiarisation, supported more comprehensive reporting of recovery, and was easier to interpret than the conventional NRS. Particularly interesting was that clinicians described using several approaches –including extra anchors—to manage uncertain interpretation of the lower numbers on the conventional NRS. This uncertainty was not deemed a problem for the SPARS. These adaptations required extra time and effort from clinicians, including to explain the anchor adaptations to external stakeholders. Considering that unplanned anchor adaptations are likely to compromise inter-rater reliability, the clarity of the SPARS anchors may support better reliability in repeated ratings than the NRS.

Although the SPARS anchors were clear, there was mixed feedback on how easy the SPARS was to explain to a patient, mostly because the new structure of the SPARS required an adjustment period. However, clinicians described the adjustment as rapid. Some gave suggestions for poster-style visual aids and electronic tools that could support explanation and familiarisation, and possibly even allow patients to independently provide ratings on the SPARS before consultations.

The SPARS was good at capturing change produced by an intervention expected to alter pain: the ‘ES II’ measure indicated excellent internal responsiveness at 0.9 (95% CI: 0.75 - 1.10). For the follow-up condition only—when symptoms were most likely to be improving—patient ratings on the SPARS covered more than double the range of the transformed NRS. This aligns with the qualitative feedback that the SPARS allowed reporting of “the feeling that there was still some improvement to go even though they weren’t really having pain anymore”, indicating the superior perceptual scope conferred by the opportunity to provide ratings within a non-painful range, particularly as a person recovers from a painful condition. The comparative ‘floor effect’ imposed by the ‘no pain’ anchor on the conventional NRS was also seen in previously published laboratory testing of the SPARS, which used laser stimuli to elicit non-painful and painful percepts in healthy volunteers (Madden et al., 2019). The current study now reveals the implications of the NRS’s floor effect for clinical utility: the SPARS avoids the interpretational difficulties that result from idiosyncratic interpretation or adding of anchors to the NRS, and provides a valued opportunity for rating of percepts that are not painful but not yet ‘right’.

After using the SPARS in this study, several clinicians reported a preference for the SPARS over the NRS, either individually or as a team. Reasons for this included ease of use and interpretation, and that the non-painful range of the SPARS reminded both patient and clinician that treatment goals go beyond pain reduction. Clinicians pointed out that many patients still wish to report stiffness or other discomfort that does not qualify as ‘pain’ but still warrants clinical attention. A different potential application for graded reporting of non-painful events would be for side-to-side comparisons of provocation tests, such as palpation, joint or soft tissue mobilisations, or end-of-joint-range assessments. For example, deep palpation of a ligament can be quite uncomfortable—approaching the pain threshold on the SPARS—in a non-painful or ‘unaffected’ joint, but frankly painful on the ‘affected’ side. Here, allowing the patient to report an ‘almost-painful’ experience in an unaffected joint simultaneously avoids misinterpretation of the report as painful (if rated NRS>0) and provides feedback to the patient that almost-painful discomfort to palpation of a ligament is normal. Although we did not specifically compare the SPARS to the NRS for provocation tests, we would anticipate that it would perform well for this purpose. Despite these advantages, enthusiastic uptake of the SPARS is likely to be tempered by the requirements of some standardised reporting (e.g. for disability or insurance assessments) that the conventional NRS be used to report pain.

Patient-reported outcome measures should perform well on the criteria of face validity, responsiveness to change, discriminating power, and test-retest reliability – although a trade-off between these criteria may be necessary. The current study addressed face validity and responsiveness to change. Participants expressed satisfaction with the opportunity for self-expression provided by the SPARS, although qualitative data collected directly from the (patient) raters themselves would be even more convincing in this regard. Responsiveness to change was excellent. It would now be helpful to clarify clinical test-retest reliability and minimum clinically meaningful change on the SPARS. With respect to test-retest reliability, the range of the SPARS may be influential. When used as a discrete scale, the SPARS offers 101 response options, and even more if used as a continuous scale. Some studies have reported that scales with more than 7-9 points offer too many response options for users to meaningfully distinguish between isolated stimuli, compromising test-retest reliability (Miller, 1994). Our own previous data showed considerable variance in within-participant ratings of experimental stimuli on both the SPARS and on a 101-point NRS (Kamerman et al., 2018; Madden et al., 2019; Madden et al., 2021). However, the 7-9-option threshold for meaningful differentiation may apply only to unidimensional stimuli: studies of multidimensional experiences suggest a capacity to meaningfully distinguish between more response options (Euasobhon et al., 2022; Preston & Colman, 2000). Given this controversy and that clinical pain is indisputably a multidimensional experience, determining the test-retest reliability of the SPARS will be an important priority going forward.

With respect to the amount of change in SPARS rating that reflects a clinically meaningful change, we are cautiously optimistic. Although Rasch-based data on the conventional NRS suggest that minimal clinically important difference varies across the NRS range (Walton, Elliott, Salim, & Al-Nasri, 2018), our experimental data showing a curvilinear stimulus-response curve for the SPARS (Madden et al., 2019), in contrast to the established power stimulus-response curve for the NRS, suggest that this weakness may not be true of the SPARS. Formal testing of this is an important focus for further study.

The original rationale for developing the SPARS was to give respondents an opportunity to provide graded reporting on non-painful events. The current data support the importance of this for the clinical context. These findings are particularly timely, given the ongoing work to develop patient-reported outcome measures using a broadly consultative, rigorous process that incorporates perspectives from people with lived experience of pain (Langford et al., 2022). It is worth noting that that ongoing work seems to be limited to adapting existing legacy measures (thus constraining ratings within a range that grades painfulness) and, in its first phase, used a cross-sectional design to obtain data from patients with actively painful conditions. That study design would be unlikely to reveal any potential need for graded reporting on non-painful but clinically relevant percepts. In contrast, the current study’s experimental-and-longitudinal design allowed for the possibility that actively painful conditions resolved into non-painful percepts during the study, and, perhaps unsurprisingly, found considerable support for a scale range that allows reporting of non-painful percepts, particularly for the later stages of recovery. Indeed, one clinician emphasised that a lower scale anchor resembling “completely normal” would be ideal for the clinical context. It is possible that this non-painful range is important only in the context of rehabilitation, but it seems more likely that it would be similarly valuable in other contexts of recovery, such as the peri-operative hospital context, primary health care, and return-to-work programmes. The current study’s mixed methods approach was particularly helpful in that the qualitative data could confirm, contradict, or extend the impressions provided by the quantitative data, and this added nuance was useful in interpreting the data. Given these benefits, we would support the continued use of mixed methods for future clinical testing of the SPARS, ideally in varied clinical contexts in which resolution of pain is anticipated.

The current data are subject to three important limitations. First, the clinicians for the current study were personally known to author GLM, introducing a possible response bias in the service of social support. Future work would benefit from recruiting participants without an existing relationship with a study leader. Second, we tested the SPARS in a context where most participants would have been first-language English-speakers, although we did not collect language data from our participants. The original SPARS anchors were developed for first-language English-speaking users; adaptation of these anchors will be necessary for other groups, may influence the utility of the adapted scale, and will require careful testing. Third, the relative effectiveness of the ‘sham’ and ‘active’ interventions was likely inconsistent across the sample, due to the pragmatic approach of allowing clinicians to select the interventions using their own discretion. However, the active intervention was likely better targeted to each patient’s complaint than the sham intervention.

Overall, this study found that the SPARS performed well and was deemed relatively easy to use and interpret. Clinicians found that it addressed important shortfalls of the conventional NRS. The strong endorsement of the scale’s structure by participating clinicians provides good rationale for further testing of the SPARS in different clinical contexts, including quantification of test-retest reliability and minimal clinically important difference.

## Data Availability

All data produced in the present work are contained in the manuscript

## Acknowledgements

Financial support: VJM is supported by the US National Institutes of Health on grant K43 TW011442. GLM and HBL are supported by an Australian NHMRC Investigator Grant awarded to G. Lorimer Moseley (ID 1178444).

Disclosures: VJM receives payment for lectures on pain and rehabilitation. PK receives consultancy fees from Partners in Research, South Africa, and is the sole proprietor of Blueprint Analytics. HBL has received speakers fees for lectures on pain and rehabilitation. LCH has received consultancy fees from Blue Note Therapeutics. GLM has received support from: Reality Health, ConnectHealth UK, Institutes of Health California, AIA Australia, Workers’ Compensation Boards and professional sporting organisations in Australia, Europe, South and North America. Professional and scientific bodies have reimbursed him for travel costs related to presentation of research on pain and pain education at scientific conferences/symposia. He has received speaker fees for lectures on pain, pain education and rehabilitation. He receives royalties for books on pain and pain education. He is non-paid CEO of the non-profit Pain Revolution and an unpaid Director of Painaustralia. The authors declare no other conflicts of interest.

## Notes

### Competing Interest Statement

The authors have declared no competing interest.

### Clinical Protocols

http://tinyurl.com/spars-peerreview

### Author Declarations

Human Research Ethics Committee of the University of South Australia gave ethics approval for this work.

### Summary of Updates

Title page: updated the number of pages in the manuscript. Revisions were made to the text, to provide clarity and reduce redundancy. Results section, Qualitative data: direct quotes in text reduced. Full raw qualitative data remain available in analysis file at OSF link. Figure 5: estimates of regression line slope and its 95% CI embedded into plots.

