## Supplementary files for "The Sensation and Pain Rating Scale: easy to use, clear to interpret, and responsive to clinical change"

### Supplementary file

#### Supplementary information on methods

##### Sample size and power calculations

Sample size was determined on the basis of precision estimates of a representative distribution of SPARS ratings taken from a pilot trial in a clinical setting, and on extrapolation of published accounts of SPARS data obtained in the experimental context [6, 9, 10]. For this experiment, the analysis requiring the largest sample was that to determine whether the SPARS yields different ratings from the NRS for percepts within the painful range. We used clinical pilot data obtained by one clinician (GLM), who administered the NRS and the SPARS with the question 'What was your average pain over the last two days?' Data were collected from 20 consecutive patients who presented for care with a chronic pain disorder. Using an online sample size calculator (HyLown.com), mean NRS ratings, mean SPARS ratings/5 (to convert the 0 to +50 range of the SPARS to a rating out of 10), a standard deviation across both of 1.5, an acceptable difference of 0.1, 80% power and alpha of 0.05, we required 87 NRS and SPARS rating pairs. We allowed for 10% missing data, which meant we aimed recruit 96 patient participants for Phase 1. We aimed to recruit 25 patients for Phase 2, in which patients completed only the SPARS (to verify that the utility and integrity of ratings on the SPARS are not dependent on, or confounded by, also obtaining ratings on the NRS).

##### Questions administered electronically

How easy or difficult was it to explain the SPARS to your patients? (Likert scale)

1. How easy or difficult was it to explain the NRS to your patients? (Likert scale)
2. Please add any comments on this AND tell us how we could make it easier for you to explain the SPARS and/or the NRS. (Free text)
3. How easy or difficult was it for your patients to report a SPARS rating? (Likert scale)
4. How easy or difficult was it for your patients to report a NRS rating? (Likert scale)
5. Please add any comments on this AND tell us how we could make it easier for them to use the SPARS and/or the NRS. (Free text)
6. How easy or difficult was it for you to interpret your patients' SPARS ratings? (Likert scale)
7. How easy or difficult was it for you to interpret your patients' NRS ratings? (Likert scale)
8. Please comment on how your confidence in, and competence in, using the SPARS changed over time. (Free text)
9. Please add any comments on this AND tell us how we could make interpretation of the SPARS and/or the NRS easier for you. (Free text)
10. Please compare the SPARS to the NRS in any ways you think are relevant to a clinician working in the context in which you work. (Free text)
11. How likely are you to use the SPARS in your clinical practice from here on? (Free text)
12. How likely are you to recommend the SPARS to other clinicians? (Free text)
13. Please tell us anything else you think we should know about the SPARS, this study or anything at all really. (Free text)

| Table S1: Initial study plans (as locked into protocol) and changes made | | |
|---|---|---|
| **Research question/topic** | **Quantitative plan** | **Qualitative data** |
| | **Blinding plans** | |
| Blinding of analyst | Initial plan: Data analyst will be blinded to the order in which assessments were undertaken and the order of the active and sham intervention.<br><br>Revised plan: Given the structure of the data, implementation of the initial plan for analyst blinding was not possible. Therefore, analysts were unblinded. | |
| | **Properties of the SPARS** | |
| Ease of explanation | Present Likert scale ratings | Yes |
| Ease of reporting | Present Likert scale ratings | Yes |
| Ease of interpreting | Present Likert scale ratings | Yes |
| Is the SPARS sensitive to change in pain induced by a therapeutic intervention? | Initial plan: Regression analysis using change in rating (from before to after intervention) as the dependent variable, intervention (Active or Sham) and scale (SPARS or NRS) as fixed effects, and allowing a random intercept for individual. Of pre-specified primary interest was a main effect of intervention within the SPARS data; additional interest was in any interaction between intervention and scale that could indicate a difference in sensitivity to change between the two scales.<br><br>Concern: Regression assesses external responsiveness by using the NRS as a benchmark for the SPARS. However, given the different anchors for the two scales, the NRS is a poor benchmark for the SPARS, making external responsiveness of limited relevance.<br><br>Revised plan: Visualise data, and calculate 'effective size II' metric to estimate SPARS ability to capture change (internal responsiveness) in response to an intervention. | - |
| **Comparison between SPARS and NRS** | | |
| Clinical utility | - | Yes |
| Relative coverage of perceptual range:<br>Does the range of SPARS ratings resemble or exceed the range of NRS ratings? | Multiply NRS ratings by 10 and compare the ranges of SPARS and NRS ratings, by condition. | - |
| Agreement between ratings | Divide SPARS ratings that lie between 0 and 50 (i.e. within the range that the NRS is expected to capture) by 5, and assess agreement between converted SPARS ratings and the matching NRS ratings, using Bland-Altman plots and Lin's concordance correlation coefficient. | Clinicians interested in potential to 'convert' scores from SPARS to NRS. |
| How are events that are rated below 0 on the SPARS (i.e. in the non-painful range) rated on the NRS? | Select only the events that were rated below 0 on the SPARS and plot the SPARS and NRS ratings for those events, to visualise usage of NRS for non-painful events. | |
| **Measurement confounding** | | |
| Are SPARS ratings affected by also collecting NRS ratings? | Compare central tendency and spread of SPARS ratings between Phase 1 and Phase 2. Visually compare the same, broken down by condition. | |
| Compare with experimental context performance | Discussion | |

*Table S2: Patient information by treating clinician*

| | 1 (N=23) | 2 (N=21) | 3 (N=16) | 4 (N=19) | 5 (N=15) | 6 (N=13) | 7 (N=4) | 8 (N=7) | 9 (N=3) | Total (N=121) |
|---|---|---|---|---|---|---|---|---|---|---|
| **Gender** | | | | | | | | | | |
| Female | 17 (73.9%) | 10 (47.6%) | 7 (43.8%) | 7 (36.8%) | 4 (26.7%) | 4 (30.8%) | 4 (100.0%) | 6 (85.7%) | 3 (100.0%) | 62 (51.2%) |
| Male | 5 (21.7%) | 11 (52.4%) | 9 (56.2%) | 10 (52.6%) | 10 (66.7%) | 9 (69.2%) | 0 (0.0%) | 1 (14.3%) | 0 (0.0%) | 55 (45.5%) |
| Other | 1 (4.3%) | 0 (0.0%) | 0 (0.0%) | 2 (10.5%) | 1 (6.7%) | 0 (0.0%) | 0 (0.0%) | 0 (0.0%) | 0 (0.0%) | 4 (3.3%) |
| **Age (yrs)** | | | | | | | | | | |
| Mean (SD) | 41 (8) | 38 (10) | 43 (12) | 42 (10) | 40 (12) | 41 (7) | 39 (5) | 43 (6) | 44 (7) | 41 (10) |
| Range | 25 - 55 | 18 - 55 | 22 - 64 | 29 - 64 | 19 - 66 | 31 - 52 | 35 - 46 | 35 - 52 | 37 - 51 | 18 - 66 |
| **Nature of diagnosis** | | | | | | | | | | |
| CRPS | 3 (13.0%) | 2 (9.5%) | 1 (6.2%) | 0 (0.0%) | 0 (0.0%) | 1 (7.7%) | 1 (25.0%) | 0 (0.0%) | 1 (33.3%) | 9 (7.4%) |
| Limb | 5 (21.7%) | 6 (28.6%) | 4 (25.0%) | 4 (21.1%) | 2 (13.3%) | 4 (30.8%) | 2 (50.0%) | 2 (28.6%) | 1 (33.3%) | 30 (24.8%) |
| Pelvic | 1 (4.3%) | 1 (4.8%) | 1 (6.2%) | 1 (5.3%) | 2 (13.3%) | 1 (7.7%) | 0 (0.0%) | 2 (28.6%) | 0 (0.0%) | 9 (7.4%) |
| Spinal | 10 (43.5%) | 10 (47.6%) | 4 (25.0%) | 10 (52.6%) | 7 (46.7%) | 4 (30.8%) | 1 (25.0%) | 3 (42.9%) | 1 (33.3%) | 50 (41.3%) |
| Widespread | 4 (17.4%) | 2 (9.5%) | 6 (37.5%) | 4 (21.1%) | 4 (26.7%) | 3 (23.1%) | 0 (0.0%) | 0 (0.0%) | 0 (0.0%) | 23 (19.0%) |

*Table S3: Patient ratings by ongoing treatment need (p-values uncorrected; exploratory testing)*

| | no (N=44) | yes (N=77) | p value |
|---|---|---|---|
| **Duration to follow-up** | | | 0.047 |
| Mean (SD) | 28 (11) | 34 (16) | |
| Range | 5 - 59 | 5 - 62 | |
| **1-week average at follow-up (SPARS)** | | | < 0.001 |
| Mean (SD) | -34 (17) | -8 (20) | |
| Range | -50 - 3 | -46 - 32 | |
| **1-week average at follow-up (NRS)** | | | < 0.001 |
| N missing | 9 | 15 | |
| Mean (SD) | 0 (0) | 1 (1) | |
| Range | 0 - 2 | 0 - 4 | |
| **2-day average at baseline (SPARS)** | | | < 0.001 |
| Mean (SD) | 7 (14) | 26 (12) | |
| Range | -17 - 38 | -4 - 44 | |
| **2-day average at baseline (NRS)** | | | < 0.001 |
| N missing | 9 | 15 | |
| Mean (SD) | 2 (2) | 5 (2) | |
| Range | 0 - 6 | 1 - 7 | |
| **Change in SPARS from baseline to follow-up** | | | < 0.001 |
| Mean (SD) | -41 (6) | -34 (9) | |
| Range | -55 - -29 | -52 - -12 | |
| **Change in NRS from baseline to follow-up** | | | < 0.001 |
| N missing | 9 | 15 | |
| Mean (SD) | -2 (2) | -4 (1) | |
| Range | -5 - 0 | -5 - -1 | |

*Table S4: Patient information by completed recovery*

| | no (N=107) | yes (N=14) | p value |
|---|---|---|---|
| **Duration to follow-up** | | | 0.110 |
| Mean (SD) | 32 (15) | 26 (4) | |
| Range | 5 - 62 | 19 - 33 | |
| **1-week average at follow-up (SPARS)** | | | < 0.001 |
| Mean (SD) | -13 (21) | -48 (3) | |
| Range | -48 - 32 | -50 - -42 | |
| **1-week average at follow-up (NRS)** | | | 0.009 |
| N missing | 21 | 3 | |
| Mean (SD) | 1 (1) | 0 (0) | |
| Range | 0 - 4 | 0 - 0 | |
| **2-day average at baseline (SPARS)** | | | < 0.001 |
| Mean (SD) | 22 (13) | -4 (8) | |
| Range | -10 - 44 | -17 - 9 | |
| **2-day average at baseline (NRS)** | | | < 0.001 |
| N missing | 21 | 3 | |
| Mean (SD) | 4 (2) | 0 (1) | |
| Range | 0 - 7 | 0 - 2 | |
| **Change in SPARS from baseline to follow-up** | | | < 0.001 |
| Mean (SD) | -35 (9) | -44 (6) | |
| Range | -55 - -12 | -51 - -33 | |
| **Change in NRS from baseline to follow-up** | | | < 0.001 |
| N missing | 21 | 3 | |
| Mean (SD) | -3 (1) | -0 (1) | |
| Range | -5 - 0 | -2 - 0 | |
